# IGI-LuNER: single-well multiplexed RT-qPCR test for SARS-CoV-2

**DOI:** 10.1101/2020.12.10.20247338

**Authors:** Elizabeth C. Stahl, Connor A. Tsuchida, Jennifer R. Hamilton, Enrique Lin-Shiao, Shana L. McDevitt, Erica A. Moehle, Lea B. Witkowsky, C. Kimberly Tsui, Kathleen Pestal, Holly K. Gildea, Matthew McElroy, Amanda Keller, Iman Sylvain, Clara Williams, Ariana Hirsh, Alison Ciling, Alexander J. Ehrenberg, IGI SARS-CoV-2 consortium, Fyodor D. Urnov, Bradley R. Ringeisen, Petros Giannikopoulos, Jennifer A. Doudna

## Abstract

Commonly used RT-qPCR-based SARS-CoV-2 diagnostics require 2-3 separate reactions or rely on detection of a single viral target, adding time and cost or risk of false-negative results. Currently, no test combines detection of widely used SARS-CoV-2 E- and N-gene targets and a sample control in a single, multiplexed reaction. We developed the IGI-LuNER RT-qPCR assay using the <u>Lu</u>na Probe Universal One-Step RT-qPCR master mix with publicly available primers and probes to detect SARS-CoV-2 N gene, E gene, and human RNase P (<u>NER</u>). This combined, cost-effective test can be performed in 384-well plates with detection sensitivity suitable for clinical reporting, and will aid in future sample pooling efforts, thus improving throughput of SARS-CoV-2 detection.

**Graphical Abstract:** 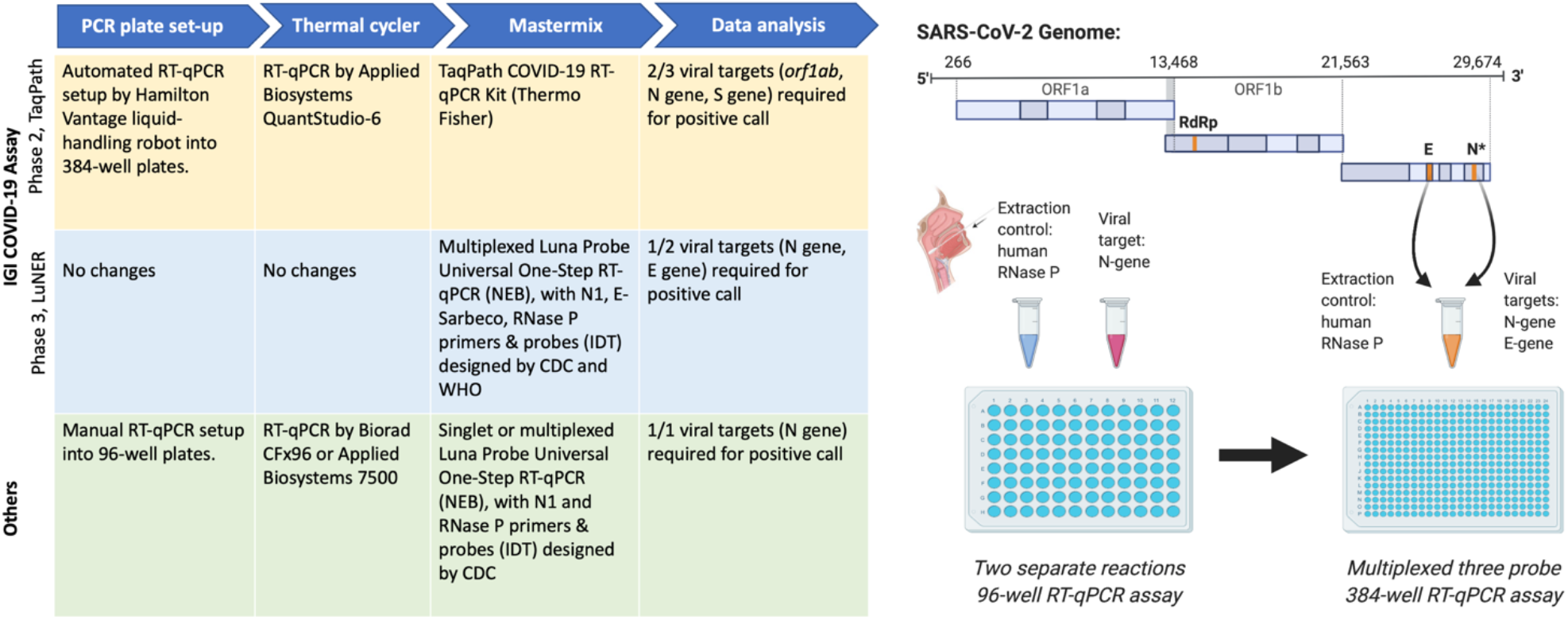

## Introduction

Late in 2019, a novel coronavirus termed SARS-CoV-2 emerged in Wuhan, China and quickly spread across the globe, prompting shelter-in-place and social distancing mandates^1–3^. One year later, over 63.5 million infections and 1.4 million deaths have been attributed to the virus globally^4^. A vital component of the public health response to slow the spread of the SARS-CoV-2 is diagnostic testing to identify positive patients, followed by isolation and contact tracing^5^. Quantitative reverse transcription polymerase chain reaction (RT-qPCR) of RNA extracted from nasal swabs is the most common diagnostic testing method to detect SARS-CoV-2^6,7^.

Current tests rely on the detection of genomic or sub-genomic viral RNA corresponding to the envelope (E) gene, the nucleocapsid (N) gene, the spike (S) gene, and/or the *ORF1ab* locus^8–10^, as well as suitable controls. However, variability in test format as well as reagent availability, quality, and cost continue to hamper widespread use for both symptomatic and asymptomatic testing. One commercially available test entered the scene early, allowing clinical laboratories to rapidly detect multiple viral genes and control RNA in a single reaction; however, this test has reported risks of yielding inaccurate results^11^ and suffers from supply shortages and high cost. Another less-expensive test detects only a single viral locus, which can lead to false-negative results^12,13^ and several tests require 2-3 separate PCR reactions to yield clinical results^9,14,15^. To address these shortcomings, the IGI Clinical Laboratory^16^ developed a new multiplexed assay with optimized viral and control RNA detection using inexpensive, widely available reagents.

An ideal test would offer affordable and robust detection of multiple SARS-CoV-2 genes and a sample extraction control using a single-well multiplexed format. We began our experiments with the Saliva Direct assay^12^, which utilizes the CDC’s published PCR primers and probes to detect the SARS-CoV-2 N-gene (N1) and human Ribonuclease P (RNase P) in a 96-well format. We then adapted the test to work in a 384-well format and to accurately detect viral RNA from either saliva or nasal-swab samples. Finally, we added a third target, SARS-CoV-2 E gene,^10^ and demonstrated that N gene, E gene, and RNase P could be multiplexed with greater than 95% clinical concordance. This high-throughput and affordable assay is demonstrated to provide sensitive detection of SARS-CoV-2 in clinical samples.

## Results

TaqMan-based RT-qPCR relies on the amplification of a reverse-transcribed product quantified in real-time by release of a fluorescent dye from a probe. By labeling different probes with spectrally distinct fluorescent dyes, such as FAM, VIC/HEX/SUN, and Cy5/ATTO647, PCR reactions can be multiplexed to detect three different targets in a single reaction (Table 1).

**Table 1.**
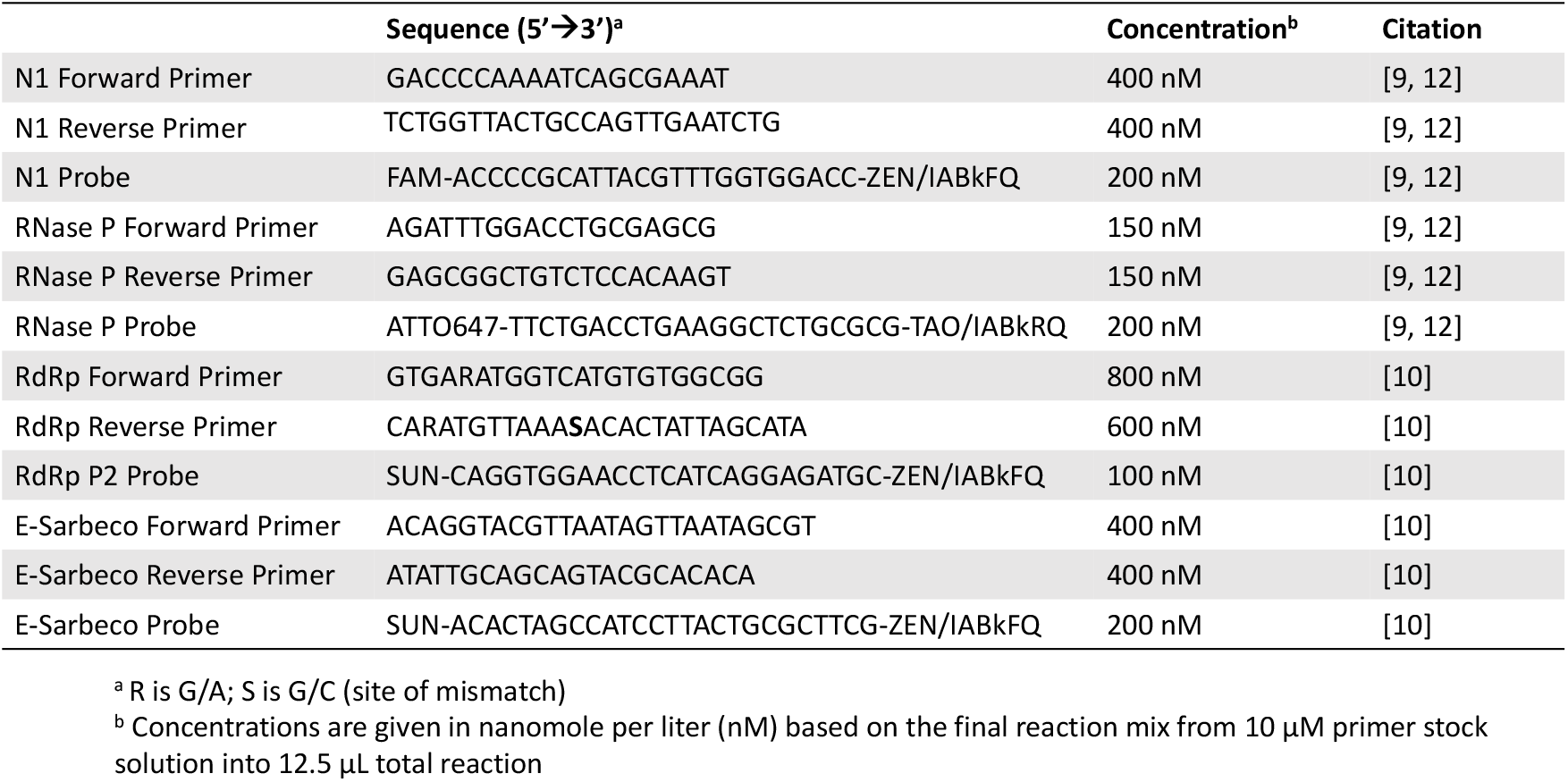
Primer and probes tested for the development of the IGI-LuNER assay.

To establish published duplex reactions^12^ (i.e. two distinct PCR targets per well) in our facility, RT-qPCR was first performed on contrived samples for N1 and RNase P using the Bio-Rad CFx96. Amplification of N1 and RNase P was observed in duplexed reactions using either full (20μL) or half-reaction volumes (Figure1A). Additionally, the duplexed reactions were successfully adapted to the Applied Biosystems QuantStudio-6 using a 384-well plate format after custom dye calibration to detect ATTO 647 (Figure 1B-C), with both the standard 2x and a newly developed 4x Luna Probe One-Step RT-qPCR master mix. This more concentrated 4x master mix contains a passive reference dye that is compatible across many instruments as well as dUTP to prevent carryover of contaminants and allows for larger sample input and multiplexing.

**Figure 1.**
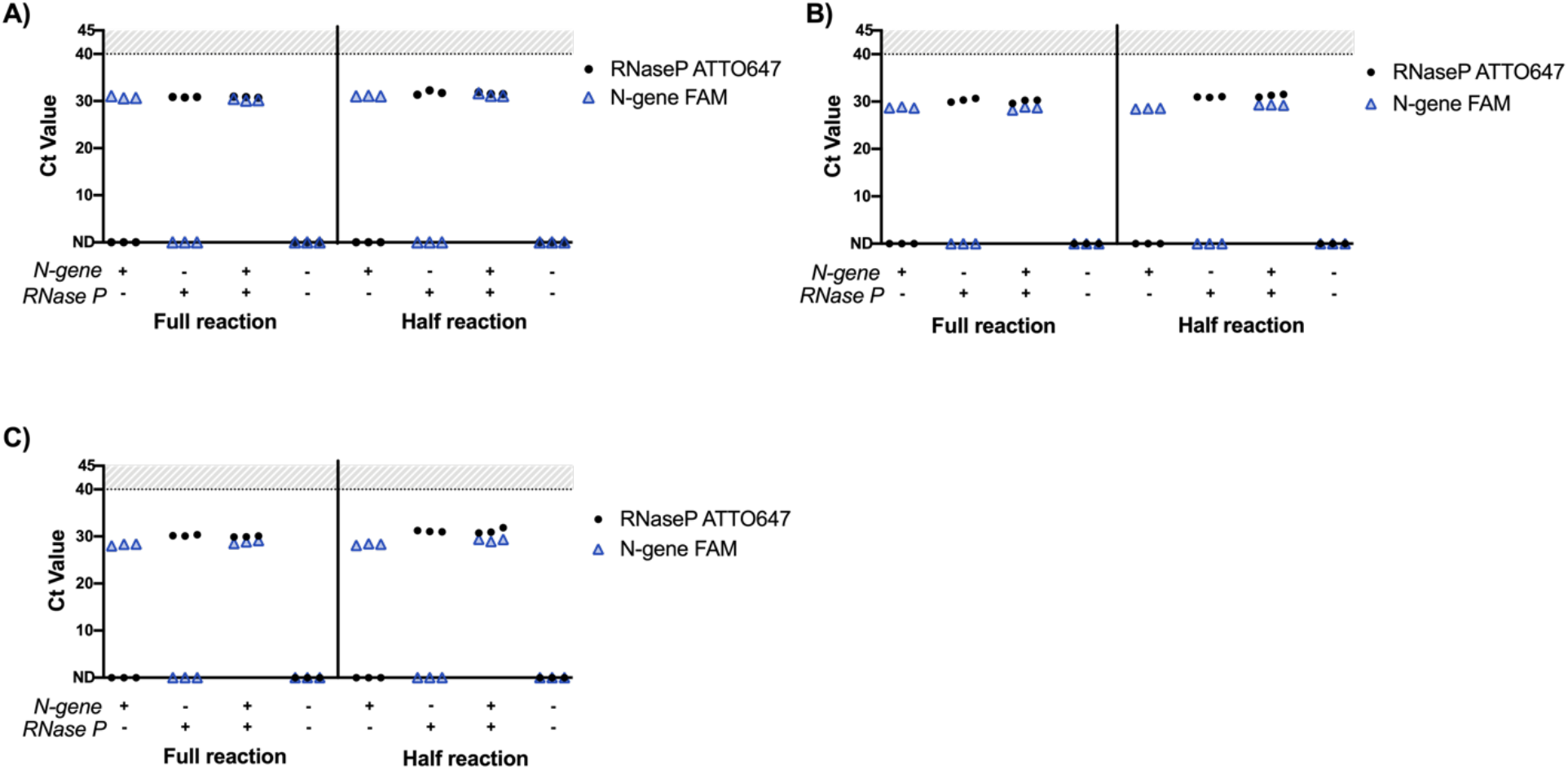
Implementation and adaptation of the Saliva Direct RT-qPCR assay to detect SARS-CoV-2 N1 and human RNase P with the QuantStudio-6. Plasmid DNA was added directly into the master mix at 200,000 copies/mL (2-4x Saliva Direct limit of detection) in triplicate. A) Full and half reaction volumes on the CFx96 with 2x master mix, B) Full and half reaction volumes on the QuantStudio-6 with 2x master mix, D) Full and half reaction volumes on the QuantStudio-6 with 4x master mix. Samples which failed to amplify are denoted as “not detected” (ND).

There was no change in the average Ct values between the 2x and 4x master mix formulations on the QuantStudio-6; however, Ct values for N1 increased by an average of 0.5 and RNase P by 1 in the duplexed, half-reaction volume compared to full reaction volume. There was no observed background amplification in the reaction without the addition of template DNA. Taken together, these data establish that duplexed reactions to detect N1 and RNase P can be adapted to 384-plate format and alternate instrumentation with minimal loss of sensitivity, while allowing for four-times greater sample throughput.

Using a single target to detect the presence of SARS-CoV-2 in a specimen may increase the risk for false-negative results^13^. One way to improve this is by integrating a second viral target into the assay, such as the N2 nucleocapsid locus^17^; however, published data show that N2 is less sensitive than N1^14^. To diversify the viral RNA regions sampled in the IGI RT-qPCR assay, both E gene and the RNA-dependent RNA polymerase (RdRp) locus of *ORF1ab* were evaluated^13^. RdRp-SARSr and E-Sarbeco (Charité) were successfully combined with RNase P in duplexed, half-reactions (Figure 2A-B). However, one of the three technical replicates of RdRp-SARSr was detected at a high Ct value of 36.3, identifying potential variability with these primers and probes.

**Figure 2.**
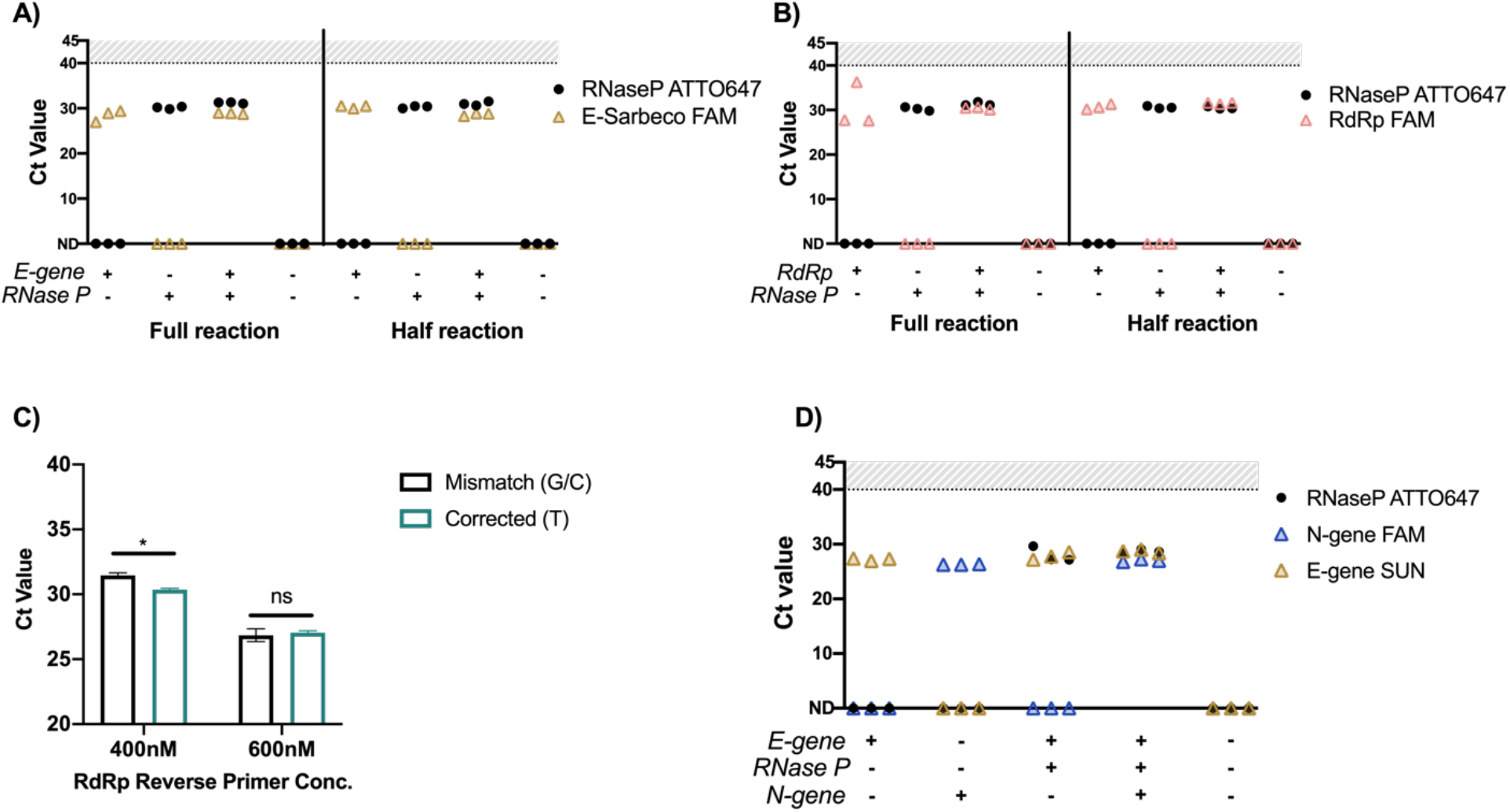
Evaluating E-Sarbeco and RdRp-SARSr primers and probes with RNase P on the QuantStudio-6. A) Full and half-sized reactions of E-Sarbeco on the QuantStudio-6 with 4x master mix B) Full and half-sized reactions of RdRp on the QuantStudio-6 with 4x master mix. Plasmid DNA was added directly into the master mix at 200,000 copies/mL (2-4x Saliva Direct limit of detection) in triplicate. C) Evaluation of corrected RdRp reverse primers at two different concentrations on pooled RNA eluted from positive clinical swab samples in triplicate. p<0.001 (*) between mismatch and corrected at 400nM and between both concentrations of primers. Not significant (ns). D) E-Sarbeco primers and probes multiplexed with N1 and RNase P in a single half-sized reaction on the QuantStudio-6 with 4x master mix.

The RdRp primers are recommended to be used at high concentrations of 600-800nM^10^ and were found to be less sensitive than other commonly used SARS-CoV-2 reagents, potentially due to a mismatch between the reverse primer and viral genome^14^. In order to control for the mismatch, we designed custom primers where G/C at position 12 was corrected to T. Correcting the mismatch improved the average Ct value by 1.1 (from Ct=31.4 to 30.3) when the reverse primer was tested at a final concentration of 400nM (p<0.01, Figure 2c). Interestingly, at the higher concentration, there was no difference in Ct values between the mismatch and corrected primers (Ct =26.8 to 27.0), suggesting the mismatch is not the primary cause of reduced sensitivity. The necessity of using high primer concentrations for the RdRp target makes it impossible to multiplex with N1 and RNase P using the standard 2x master mix. Multiplexing was achieved using the 4x master mix by increasing the stock concentrations of primers and probes. These data suggest multiplexing with existing RdRp primers and probes is challenging but can be achieved if reagents are amenable to smaller input volumes.

The E-Sarbeco primers and probes were successfully multiplexed in PCR reactions using both the 2x and 4x master mix formulations at the standard input concentration of 10μM (Figure 2D) for a final primer concentration of 400nM, demonstrating superior performance over RdRp. Therefore, we have selected the 4x <u>Lun</u>a master mix to detect <u>N</u>-gene, <u>E</u>-gene, and <u>R</u>Nase P (NER) as the final formulation of our multiplexed assay, hereafter referred to as IGI-LuNER. Validation studies using the IGI-LuNER assay were performed to define the limit of detection, demonstrate reproducibility, and determine clinical concordance.

To determine the limit of detection (LoD), heat-inactivated SARS-CoV-2, measured as fifty-percent tissue culture infective dose per milliliter (TCID_50_/ml), was spiked into negative sample matrix generated from clinically reported negative oropharyngeal (OP)/mid-turbinate swab specimens. RNA extraction was performed with the MagMax Viral/Pathogen Nucleic Acid Isolation Kit using the Hamilton Vantage liquid-handling system, as described previously^16^. Following RNA extraction, samples were analyzed with the IGI-LuNER RT-qPCR assay. At 0.1 TCID_50_/mL, E gene was detected in 2 out of 3 replicates, and N gene was undetected. At 0.5 TCID_50_/mL, E gene was measured at Ct = 37.1 ± 2.95, and N gene was measured at Ct = 33.93 ± 0.52 in all three replicates, demonstrating the lowest concentration where all samples were called positive and thus the tested limit of detection (Figure 3A).

**Figure 3.**
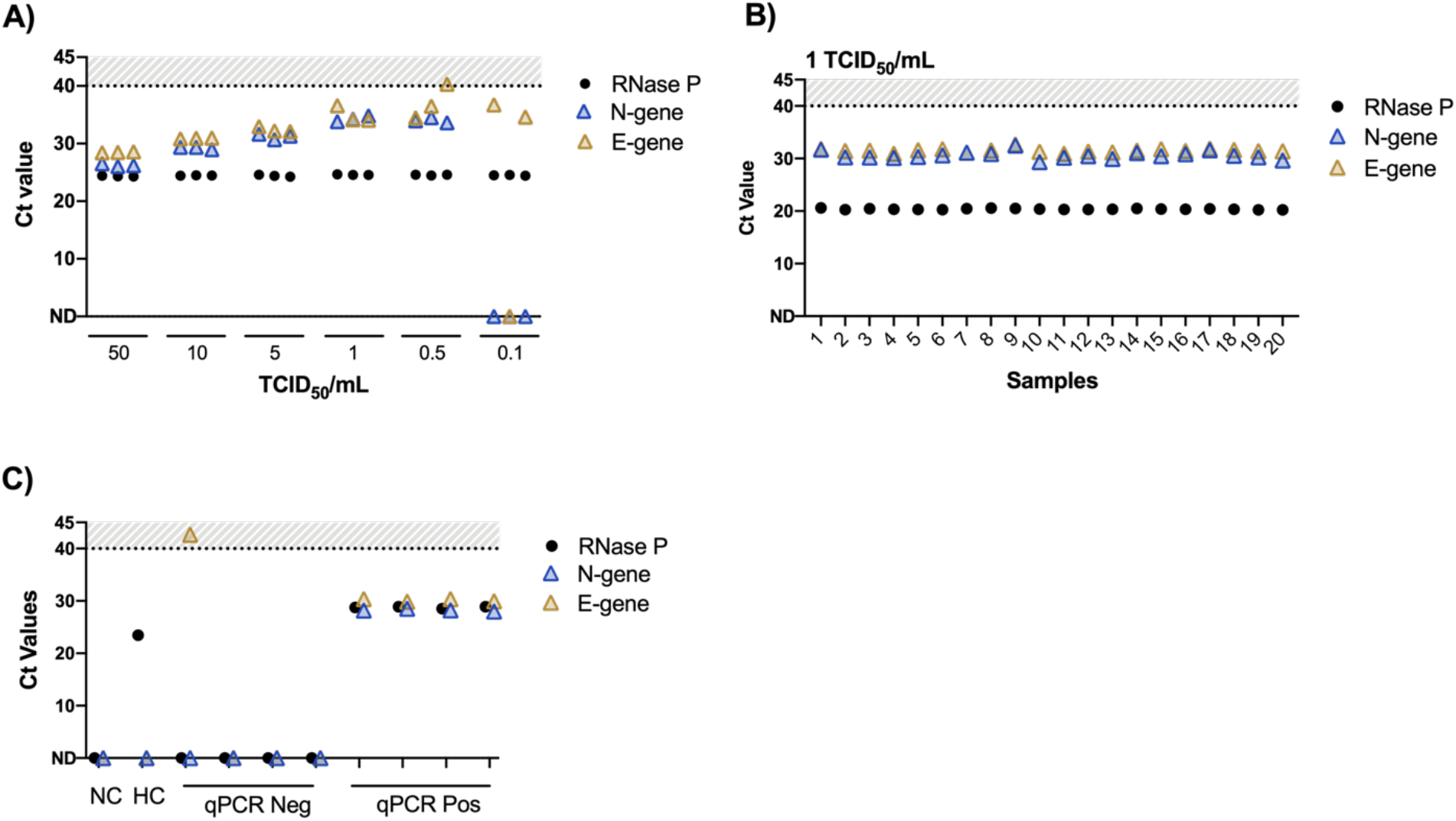
Limit of detection and reproducibility. A) The limit of detection was defined by extracting RNA from heat inactivated virus at concentrations ranging from 50 TCID_50_/mL to 0.1 TCID_50_/mL and performing RT-qPCR with the LuNER reagents. B) Twenty replicates at 2x LoD (1 TCID_50_/mL) were prepared by extracting RNA from heat inactivated virus and performing RT-qPCR with the LuNER reagents to test for reproducibility. Samples were positive if either N gene or E gene was detected at a Ct value < 40. Samples that did not amplify or had Ct values >=40 are shown on the x-axis. C) Controls for the LuNER assay are valid, including negative buffer-only control (“NC”), human RNA control (“HC”), qPCR negative, and qPCR positive controls.

In order to evaluate whether the IGI-LuNER assay would return reproducible Ct values, 20 contrived replicates were prepared using heat-inactivated virus at 2x the defined LoD in negative sample matrix. All 20 samples at 1 TCID_50_/mL were reproducibility detected (Figure 3B, average RNase P Ct = 20.4 ± 0.1, E gene Ct = 31.5 ± 0.4, N gene Ct = 30.6 ± 0.7) along with experimental controls (Figure 3C) below the Ct cutoff of 40.

Finally, to assess how the IGI-LuNER assay performs on clinical samples, 91 previously reported positive and negative samples from the IGI Clinical Laboratory were selected for a head-to-head comparison of the TaqPath RT-PCR COVID-19 kit (ThermoFisher) and the IGI-LuNER assay (Figure 4A). Internal validation criteria were defined as 95% negative agreement and 95% positive agreement. Samples were stored at 4°C for 3-15 weeks prior to RNA extraction to evaluate clinical concordance with new RT-qPCR reagents.

**Figure 4.**
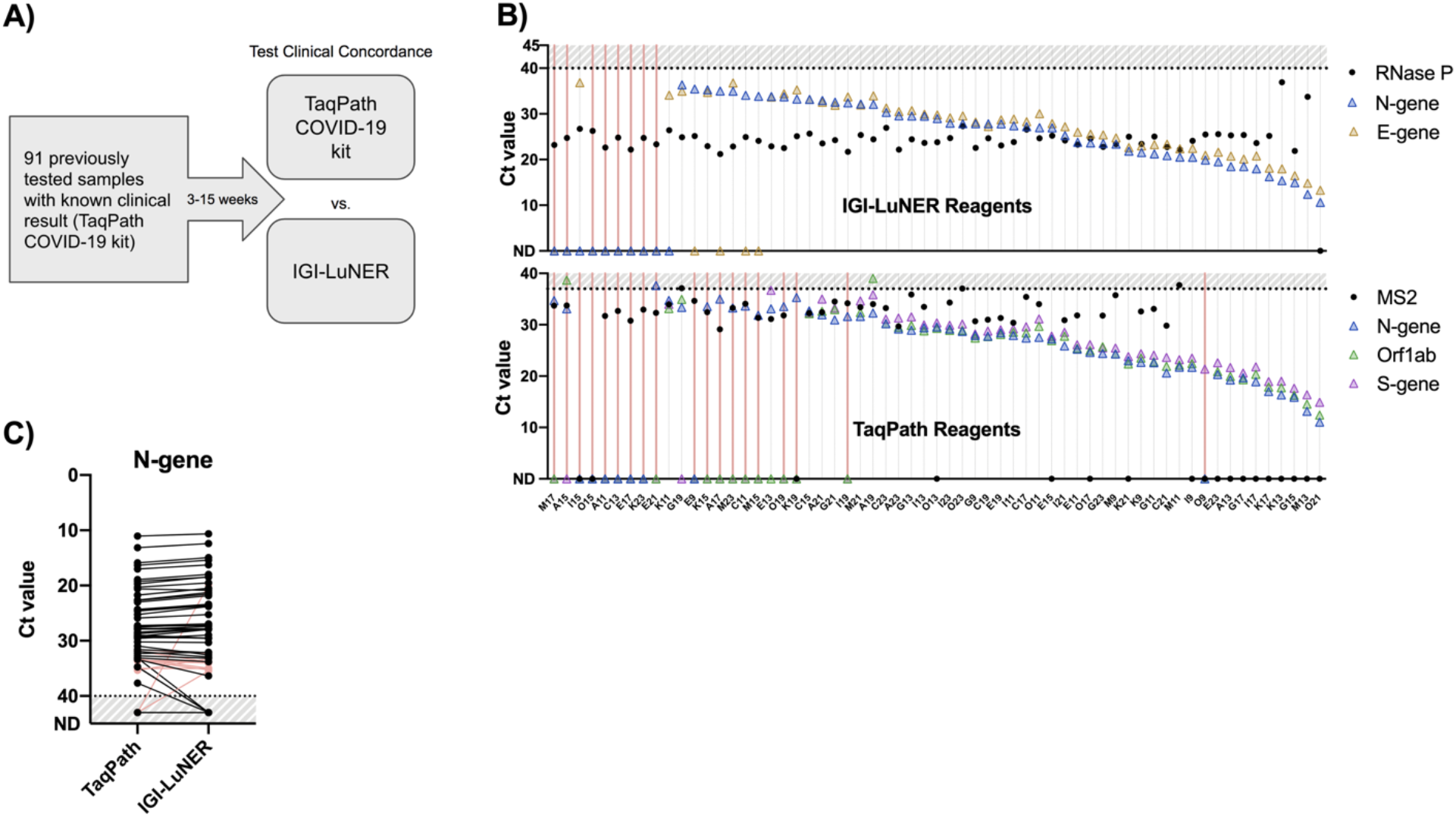
Clinical concordance. A) RNA was extracted from known clinical samples and reanalyzed with the TaqPath COVID-19 kit or IGI-LuNER assay. B) Ct value comparisons for the 61 expected positive samples show 9 negative samples in the IGI-LuNER assay (pink lines), which were also negative in the TaqPath COVID-19 kit. An additional 10 samples assayed with the TaqPath COVID-19 kit returned an alternative result. C) Ct value comparison for N-gene between the IGI-LuNER and TaqPath assays shows strong concordance. Pink lines reflect samples in the TaqPath kit that returned inconclusive or invalid results despite amplifying N-gene in the expected positive samples. Logic gating for possible sample results is listed in the Methods.

All 30 of the expected negative samples returned a negative result with the IGI-LuNER assay, with an average RNase P Ct = 24.05 ± 1.08, whereas the TaqPath RT-PCR COVID-19 kit returned 26 negative and 4 invalid results, due to failure of the extraction control, MS2 phage, to be detected (Table 2 and Supplemental Table 1). Of the 61 expected positive samples, 53 resulted positive with the IGI-LuNER assay, while 8 samples resulted as negative. The same group of 8 discordant samples returned either a negative, invalid, or inconclusive result with the TaqPath RT-PCR COVID-19 kit, along with an additional 10 samples that failed to return the expected positive result (Figure 4B). These additional non-positive samples in the TaqPath group were skewed towards higher Ct values. Comparisons of N gene, S gene, and *orf1ab* showed that samples with originally high Ct values were less likely to be detected upon retesting with both the TaqPath and IGI-LuNER assays, rather than samples that had been stored for the longest duration; and the IGI-LuNER assay demonstrated superior sensitivity to detect positive samples in this experiment (Supplemental Figures 1 and 2).

**Table 2.** Clinical evaluation of IGI-LuNER RT-qPCR with matched swab samples.

| Sample | TaqPath<br>Original | TaqPath<br>Re-Test | IGI-LuNER<br>Re-Test | Sample | TaqPath<br>Original | TaqPath<br>Re-Test | IGI-LuNER<br>Re-Test | Sample | TaqPath<br>Original | TaqPath<br>Re-Test | IGI-LuNER<br>Re-Test |
| --- | --- | --- | --- | --- | --- | --- | --- | --- | --- | --- | --- |
| A1 | NEG | NEG | NEG | A13 | POS | POS | POS | K13 | POS | POS | POS |
| A3 | NEG | NEG | NEG | A19 | POS | POS | POS | K17 | POS | POS | POS |
| A5 | NEG | NEG | NEG | A21 | POS | POS | POS | K21 | POS | POS | POS |
| A7 | NEG | NEG | NEG | A23 | POS | POS | POS | M9 | POS | POS | POS |
| C1 | NEG | NEG | NEG | C15 | POS | POS | POS | M11 | POS | POS | POS |
| C3 | NEG | NEG | NEG | C17 | POS | POS | POS | M13 | POS | POS | POS |
| C5 | NEG | NEG | NEG | C19 | POS | POS | POS | M21 | POS | POS | POS |
| C7 | NEG | INVALID | NEG | C21 | POS | POS | POS | O11 | POS | POS | POS |
| E1 | NEG | NEG | NEG | C23 | POS | POS | POS | O13 | POS | POS | POS |
| E3 | NEG | NEG | NEG | E11 | POS | POS | POS | O17 | POS | POS | POS |
| E5 | NEG | NEG | NEG | E13 | POS | POS | POS | O21 | POS | POS | POS |
| E7 | NEG | NEG | NEG | E15 | POS | POS | POS | O23 | POS | POS | POS |
| G1 | NEG | NEG | NEG | E19 | POS | POS | POS | A11 | POS | NEG | NEG |
| G3 | NEG | NEG | NEG | E23 | POS | POS | POS | C13 | POS | NEG | NEG |
| G5 | NEG | NEG | NEG | G9 | POS | POS | POS | E9 | POS | NEG | POS |
| I1 | NEG | NEG | NEG | G11 | POS | POS | POS | E17 | POS | NEG | NEG |
| I3 | NEG | NEG | NEG | G13 | POS | POS | POS | E21 | POS | NEG | NEG |
| I5 | NEG | NEG | NEG | G15 | POS | POS | POS | K23 | POS | NEG | NEG |
| I7 | NEG | NEG | NEG | G17 | POS | POS | POS | I15 | POS | INVALID | POS |
| K1 | NEG | NEG | NEG | G19 | POS | POS | POS | K19 | POS | INVALID | POS |
| K3 | NEG | INVALID | NEG | G21 | POS | POS | POS | O9 | POS | INVALID | POS |
| K5 | NEG | NEG | NEG | G23 | POS | POS | POS | O15 | POS | INVALID | NEG |
| K7 | NEG | NEG | NEG | I9 | POS | POS | POS | A15 | POS | INC. | NEG |
| M1 | NEG | NEG | NEG | I11 | POS | POS | POS | A17 | POS | INC. | POS |
| M3 | NEG | NEG | NEG | I13 | POS | POS | POS | C11 | POS | INC. | POS |
| M5 | NEG | INVALID | NEG | I17 | POS | POS | POS | I19 | POS | INC. | POS |
| M7 | NEG | NEG | NEG | I21 | POS | POS | POS | K15 | POS | INC. | POS |
| O1 | NEG | NEG | NEG | I23 | POS | POS | POS | M15 | POS | INC. | POS |
| O3 | NEG | NEG | NEG | K9 | POS | POS | POS | M17 | POS | INC. | NEG |
| O5 | NEG | INVALID | NEG | K11 | POS | POS | POS | M23 | POS | INC. | POS |
|  |  |  |  |  |  |  |  | O19 | POS | INC. | POS |

Ct value comparisons between N gene in the two assays were very similar: of the paired measurements, the IGI-LuNER assay returned a lower Ct value from 30 of the samples (Ct decreased by 0.86 ± 0.43) and greater Ct value from 16 of the samples (Ct increased by 0.84 ± 0.83, Figure 4C). In summary, the IGI-LuNER showed 100% negative agreement with the original sample results and 90% positive agreement with the original sample result, where the discordant samples were in 100% agreement when retested using the TaqPath RT-PCR COVID-19 kit, surpassing the validation criteria.

## Discussion

The 2020 SARS-CoV-2 pandemic has cast an unprecedented level of attention on technical specifications of PCR-based tests as well as their affordability and supply chain dependence. One of the drawbacks of PCR diagnostics is the lengthy turnaround time due to the complexity of molecular biology techniques involved. Performing PCR reactions in a 384-well format allows for a greater number of samples to be processed each day, resulting in shorter turnaround times, and also reduces the cost per sample by lowering reagent volumes and thus supply chain dependence. While other groups have adopted extraction-free methods, 384-well formats, or multiplexed PCR reactions to improve turnaround times, these assays are limited by the fact that they 1) target a single site in the viral genome, 2) require 2-3 separate PCR reactions, or 3) utilize proprietary primer and probe sequences that are expensive and/or not widely available^18^. The IGI-LuNER assay addresses these shortcomings by combining affordable, publicly and commercially available SARS-CoV-2 reagents into a multiplexed 384-well reaction. We show that the IGI-LuNER assay has improved sensitivity and greater than 95% clinical concordance compared to a similar multiplexed assay.

The IGI-LuNER assay is estimated to cost $1.26 per reaction, a 6-12x reduction in cost from other commercially available tests. Additionally, the IGI-LuNER assay eliminates sample “Inconclusive” results based on amplification of only one viral gene and will likely reduce the number of “Invalid” results by implementing an internal human extraction control, thereby reducing the number of samples that must be re-tested for further cost and reagent savings. Of note, the RNase P primers designed by the CDC may amplify both RNA and genomic DNA^19^; thus using a redesigned RNase P reverse primer that specifically detects RNA or incorporating DNase digestion of the sample prior to RT-qPCR may improve the test further. Although vaccines are expected to be administered in early 2021, diagnostic testing for SARS-CoV-2 will remain necessary for surveillance. The IGI-LuNER assay will improve accessibility for SARS-CoV-2 testing by reducing costs and can be easily adapted to detect new SARS-CoV-2 variants or novel pathogens in the future.

## Methods

### Optimization of multiplexed RT-qPCR assay

Experiments were performed by adding 5μL of positive control plasmid DNA encoding N-gene (2019-nCoV_N_Positive Control, IDT, Coralville, Iowa), RNase P (Hs_RPP30 Positive Control, IDT), E-gene (2019-nCoV_E Positive Control, IDT), and RdRp (2019-nCoV_RdRp (ORF1ab) Positive Control, IDT)) at 200,000 copies/mL directly into 15μL (full reactions) or 7.5μL (half reactions) of master mix in 96 or 384-well plates (ThermoFisher, Waltham, MA).

The master mix was composed of Luna Probe Universal One-Step RT-qPCR (New England Biolabs, Ipswich, MA E3006, E3007, or M3019) and combinations of N1 primers (nCOV_N1 Forward Primer and nCOV_N1 Reverse Primer, IDT), N1 (FAM) probe (nCOV_N1 Probe, IDT), E-Sarbeco primers (E_Sarbeco_F1 Forward Primer and E_Sarbeco_R2 Reverse Primer, IDT), E-Sarbeco (FAM/SUN) probe (E_Sarbeco_P1 Probe, IDT), RdRp primers (RdRP_SARSr_F2 Forward Primer and RdRP_SARSr_R1 Reverse Primer, IDT), RdRp (FAM/SUN) P2 Probe (RdRP_SARSr_P2 Probe, IDT), RNase P primers (RNase P Forward Primer and RNase P Reverse Primer, IDT), and RNase P (ATTO 647) Probe (IDT) diluted in nuclease-free water. Final concentrations of individual primers and probes are listed in Table 1. The input concentration of the primers and probes was 10μM unless otherwise noted.

The following thermal cycling conditions were used on the CFx96 (Bio-Rad, Hercules CA) and QuantStudio-6 (Applied Biosystems, Foster City, CA): 10 minutes at 52°C, 2 minutes at 95°C, 45 cycles with 10 seconds at 95°C and 30 seconds at 55°C. Data was collected for FAM, Cy5/ATTO 647 (custom dye), and VIC (SUN). Data was analyzed using the Design and Analysis software (ThermoFisher). Ct values were exported into Excel version 16.42 (Microsoft Office, Redmond WA) and plotted with Prism 8.4.3 (Graphpad, San Diego, CA) as the average value ± standard deviation.

### Acquisition and Processing of Clinical Samples

The Innovative Genomics Institute Clinical Laboratory (IGI) is a high-complexity, CLIA-certified laboratory, licensed to operate in the State of California through the California Department of Public Health. We confirm all relevant ethical guidelines have been followed, and any necessary IRB and/or ethics committee approvals have been obtained.

### RNA Extraction of Clinical Samples

Oropharyngeal (OP, Improve Medical, China) swabs were used to collect OP/mid-turbinate samples from individuals at walk-up testing centers in Berkeley, CA. Swabs were transported to the IGI Clinical Laboratory in 4mL of 2x DNA/RNA Shield (Zymo Research, Irvine CA), diluted to 1x with sterile phosphate buffered saline, pH 7.4 (Gibco, Gaithersburg, MD). Samples were kept at 4°C until 450μL was aliquoted into 96-well deep-well plates (Axygen, San Francisco CA) with the Microlab STARlet liquid-handling robot (Hamilton, Reno NV). RNA extraction was performed on the Vantage liquid handling system (Hamilton) with the MagMax Viral/Pathogen Nucleic Acid Isolation Kit (Applied Biosystems), according to the manufacturer’s instructions, with the exception of omitting MS2 spike-in for the IGI-LuNER assay. Final RNA elution volume was 22μL. RNA was stored at −80°C or arrayed into 384-well master mix plates on the Vantage liquid-handling system (5μL of sample into 7.5μL master mix) for RT-qPCR.

### Limit of Detection

Heat-inactivated virus was generated in the BSL3 laboratory at the University of California, Berkeley. Briefly, SARS-CoV-2 was cultured in Vero E6 cells and titers were determined using cytopathic effect (CPE) assays. Viral samples were incubated at 37°C for 30 minutes in the presence of 0.5 mg/mL proteinase K, followed by 75°C for 30 minutes. The starting titer was 158,000 TCID_50_/mL. To determine the limit of detection, six clinically reported negative samples (OP/mid-turbinate swabs in 4mL of 1x RNA/DNA shield) were pooled to generate a negative sample matrix for spike-in experiments. 45μL of 10x heat-inactivated virus was added to 405μL of sample matrix to generate 450μL of material for RNA extraction using the methods described above at final concentrations of 50, 10, 5, 1, 0.5, and 0.1 TCID_50_/mL in triplicate. Following RNA extraction, the samples were arrayed into the LuNER master mix as described above and RT-qPCR was performed using half-reaction volumes on the QuantStudio-6. Data was analyzed using the Design and Analysis software (ThermoFisher). Ct values were exported into Excel version 16.42 (Microsoft Office, Redmond WA) and plotted with Prism 8.4.3 (Graphpad, San Diego, CA) as the average value ± standard deviation.

### Reproducibility

Eight clinically reported negative samples (OP/mid-turbinate swabs in 4mL of 1x RNA/DNA shield) were pooled to generate a negative sample matrix for spike-in experiments. 45μL of 10x heat-inactivated virus was added to 405μL of sample matrix to generate 450μL of material for RNA extraction using the methods described above at a final concentration of 1 TCID_50_/mL with twenty-replicates each.

### Clinical Concordance

91 OP/mid-turbinate samples with clinically reported results (61 positive, 30 negative), based on original RNA extraction with the MagMax Viral/Pathogen Nucleic Acid Isolation Kit and the TaqPath RT-PCR COVID-19 kit (ThermoFisher), were arrayed from the original tubes (containing residual volume of approximately 3.5mL of 1x DNA/RNA shield) into a 96-well deep-well plate on the Microlab STARlet. The samples and extraction controls underwent a second round of RNA extraction then RT-qPCR with the TaqPath RT-PCR COVID-19 kit and IGI-LuNER assay in parallel to study percent positive and negative agreement. Samples were stored at 4°C upon arrival and up to 15 weeks before retesting.

### Sample and Control Logic and Resulting

#### IGI-LuNER

The IGI-LuNER assay was developed and its performance characteristics were determined in accordance with CLIA, specifically 42 CFR 493.1253. It has been approved for clinical use by IGI’s laboratory director and should not be regarded as investigational or for research purposes. This assay has not yet been cleared or approved by the US Food and Drug Administration (FDA), since FDA does not require that this assay go through premarket FDA review, including assays related to COVID-19.

| Sample Result | Extraction control | Viral Targets |
| --- | --- | --- |
| Positive | - | Either E-gene **or** N-gene Ct < 40 |
| Negative | RNase P Ct < 35 | Both E-gene **and** N-gene Ct ≥ 40 |
| Invalid | RNase P Ct ≥ 35 | Both E-gene **and** N-gene Ct ≥ 40 |

| Controls | Valid when |
| --- | --- |
| NC | RNase P Ct ≥ 35, both E-gene **and** N-gene Ct ≥ 40 |
| HC | RNase P Ct < 35, both E-gene **and** N-gene Ct ≥ 40 |
| qPCR Negative | RNase P Ct ≥ 35, both E-gene **and** N-gene Ct ≥ 40 |
| qPCR positive | RNase P Ct < 35, both E-gene **and** N-gene Ct < 40 |

The Ct cut-off of 40 was established by examining background amplification in known negative swab samples when the thresholds were set just above the negative qPCR control (water). Spurious amplification could be occasionally observed at Ct values > 42. A sample is called positive when N gene or E gene has Ct values < 40 (RNase P is ignored). A sample is called negative when N gene and E gene have Ct values ≥ 40 and RNase P has a Ct value < 35. A sample is called invalid when N gene and E gene have Ct values ≥ 40 and RNase P has Ct ≥ 35. Ct values may be returned as “undetermined” if there is no amplification (treated the same as Ct ≥ 40 or ≥ 35). Samples that result as invalid are re-tested from RNA extraction.

Two controls undergo the full process of RNA extraction. The “NC” control (50:50 2xRNA Shield:1xPBS) is valid when N gene and E gene have Ct values ≥ 40 and RNase P has Ct ≥ 35, controlling for background amplification or contamination. The “HC” control (50:50 2xRNA Shield:1xPBS with human RNA) is valid when N gene and E gene have Ct values ≥ 40 and RNase P has a Ct value < 35, controlling for non-specific amplification, contamination, and retention of genomic material. Prior to RT-qPCR, four wells in the 384-well plate receive nuclease-free water as “qPCR Negative Controls,” which are valid when N gene and E gene have Ct values ≥ 40 and RNase P has Ct ≥ 35. Additionally, four wells receive positive control DNA plasmid (“qPCR Positive Controls”) at 100,000 copies/mL and are valid when N gene and E gene have Ct values < 40 and RNase P has a Ct value < 35.

#### TaqPath RT-PCR

TaqPath RT-PCR:
| Sample Result | Extraction spike-in control (Bacteriophage MS2) | Viral Targets (*S-gene*, *N-gene*, *ORF1ab*) |
| --- | --- | --- |
| Positive | Not required | ≥2 of 3 viral targets have Ct < 37 |
| Negative | MS2 Ct < 37 | 0 viral targets have Ct < 37 |
| Invalid | MS2 Ct ≥ 37 | 0 viral targets have Ct < 37 |
| Inconclusive | MS2 Ct < 37 | 1 of 3 viral targets have Ct < 37 |

| Controls | Valid when |
| --- | --- |
| NC | MS2 Ct < 37, 3 of 3 viral targets Ct ≥ 37 |
| HC | MS2 Ct < 37, 3 of 3 viral targets Ct ≥ 37 |
| qPCR Negative | MS2 Ct ≥ 37, 3 of 3 viral targets Ct ≥ 37 |
| qPCR positive | MS2 Ct ≥ 37, 3 of 3 viral targets Ct < 37 |

A sample is called positive when 2 out of 3 tested viral genes have Ct values < 37 (N gene, S gene, *ORF1ab*; and MS2 extraction control is ignored). A sample is inconclusive when 1 out of 3 tested viral genes have Ct values < 37 (N gene, S gene, *ORF1ab*) and MS2 has a Ct value < 37. A sample is called negative when 3 out of 3 viral genes have Ct values ≥ 37 and MS2 has a Ct value < 37. A sample is called invalid when 3 out of 3 viral genes have Ct values ≥ 37 and MS2 has Ct ≥ 37. Ct values may be returned as “undetermined” if there is no amplification (treated the same as Ct ≥ 37). Samples that result as invalid or inconclusive are re-tested from RNA extraction.

Two controls undergo the full process of RNA extraction. The “NC” control (50:50 2xRNA Shield:1xPBS) is valid when all three viral genes have Ct values ≥ 37 and MS2 has Ct < 37, controlling for background amplification or contamination. The “HC” control (50:50 2xRNA Shield:1xPBS with human RNA) is valid when all three viral genes have Ct values ≥ 37 and MS2 has Ct < 37, controlling for background amplification or contamination. Prior to qRT-PCR, four wells in the 384-well plate receive nuclease-free water as “qPCR Negative Controls,” which are valid when all three viral genes have Ct values ≥ 37 and MS2 has Ct ≥ 37. Additionally, four wells receive Thermo SARS-CoV-2 positive control RNA and are valid when all three viral genes have Ct values < 37 and MS2 has Ct ≥ 37.

## Data Availability

All data included in this article and the supplementary files are available upon request to the corresponding authors.

## Acknowledgements

Thank you to New England Biolabs for the gift of the Luna Probe Universal One-Step 4x master mix and technical support. Thank you to IDT for technical support, including producing the custom E-gene SUN probe. Thank you to the Stanley lab at the University of California Berkeley and the BSL3 facility for generating the heat-inactivated virus. Thank you to the additional members of IGI Testing Consortium: Alexandra M. Amen, Kerrie W. Barry, John M. Boyle, Cara E. Brook, Seunga Choo, L. T. Cornmesser, David J. Dilworth, Indro Fedrigo, Skyler E. Friedline, Thomas G. W. Graham, Ralph Green, Megan L. Hochstrasser, Dirk Hockemeyer, Netravathi Krishnappa, Azra Lari, Hanqin Li, Tianlin Lu, Elijah F. Lyons, Kevin G. Mark, Lisa Argento Martell, A. Raquel O. Martins, Patrick S. Mitchell, Christine Naca, Divya Nandakumar, Elizabeth O’Brien, Derek J. Pappas, Diana L. Quach, Benjamin E. Rubin, Rohan Sachdeva, Abdullah Muhammad Syed, I-Li Tan, Amy L. Tollner, Timothy K. Turkalo, M. Bryan Warf, and Oscar N. Whitney.

## Funding

This study was funded by gifts from the Packard Foundation and the Curci Foundation to the Innovative Genomics Institute at UC Berkeley. J.R.H. is a Fellow of The Jane Coffin Childs Memorial Fund for Medical Research. K.P. is a Cancer Research Institute Irvington Fellow supported by the Cancer Research Institute. H.K.G is supported by NSF Grant # DGE175814 and NIA 1F99AG068343-01.

## Competing Interests

The Regents of the University of California have patents issued and pending for CRISPR technologies on which J.A.D. is an inventor. J.A.D. is a cofounder of Caribou Biosciences, Editas Medicine, Scribe Therapeutics, Intellia Therapeutics and Mammoth Biosciences. J.A.D. is a scientific advisory board member of Caribou Biosciences, Intellia Therapeutics, eFFECTOR Therapeutics, Scribe Therapeutics, Mammoth Biosciences, Synthego, Algen Biotechnologies, Felix Biosciences and Inari. J.A.D. is a Director at Johnson & Johnson and has research projects sponsored by Biogen, Pfizer, AppleTree Partners and Roche. F.D.U. is a co-founder of Tune Therapeutics. P.G. is a cofounder and Director at NewCo Health. P.G. is the CLIA Laboratory Director for Coral Genomics and 3DMed. The other authors declare no competing interests.

## Notes

### Author Declarations

Descriptive statistics for patient samples used in this manuscript come from de-identified datasets in accordance with human subjects protections as approved by the UC Berkeley Committee for Protection of Human Subjects (CPHS). While Institutional Review Board (IRB) approval is required for any research using human subjects clinical laboratory activities exclusively supporting CLIA-certified clinical operations do not. These activities are governed by CMS and HIPAA legislation. In order to publish our data in this manuscript as results of developing a testing workstream, we sought IRB approval through UC Berkeley CPHS. The UC Berkeley Committee for Protection of Human Subjects determined that all the analyses presented in this manuscript do not qualify as human subjects research as the data sets were de-identified to those analyzing them for these results (IRB submission # 2020-04-13177).

### Summary of Updates

Table 2 and associated text revised to correct K23 sample result

