## Supplemental Materials for "IGI-LuNER: single-well multiplexed RT-qPCR test for SARS-CoV-2"

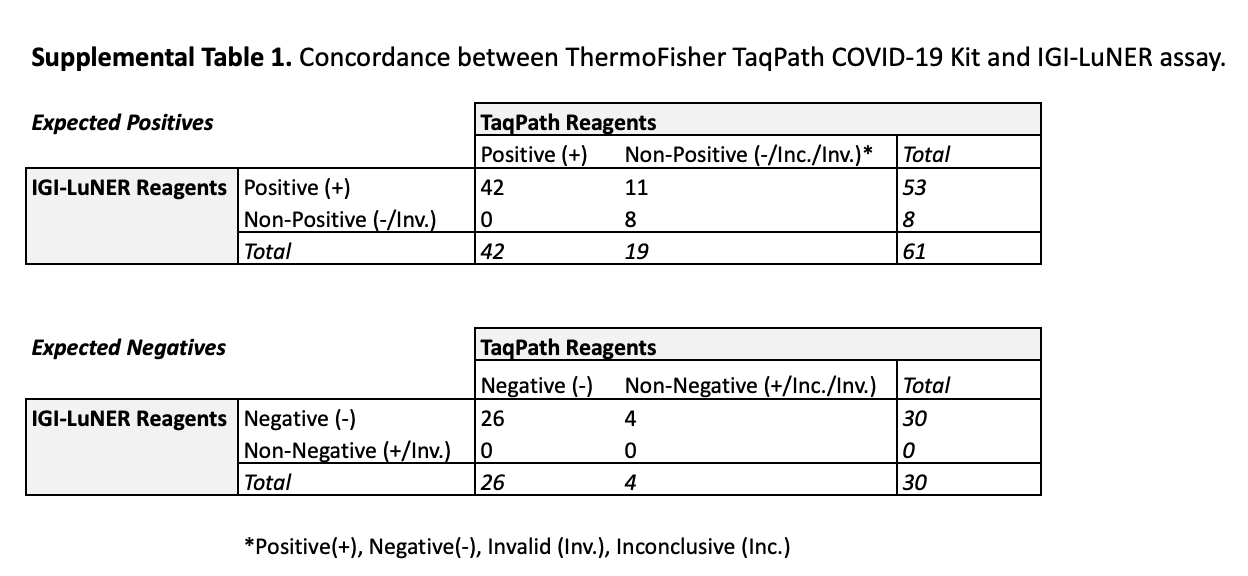

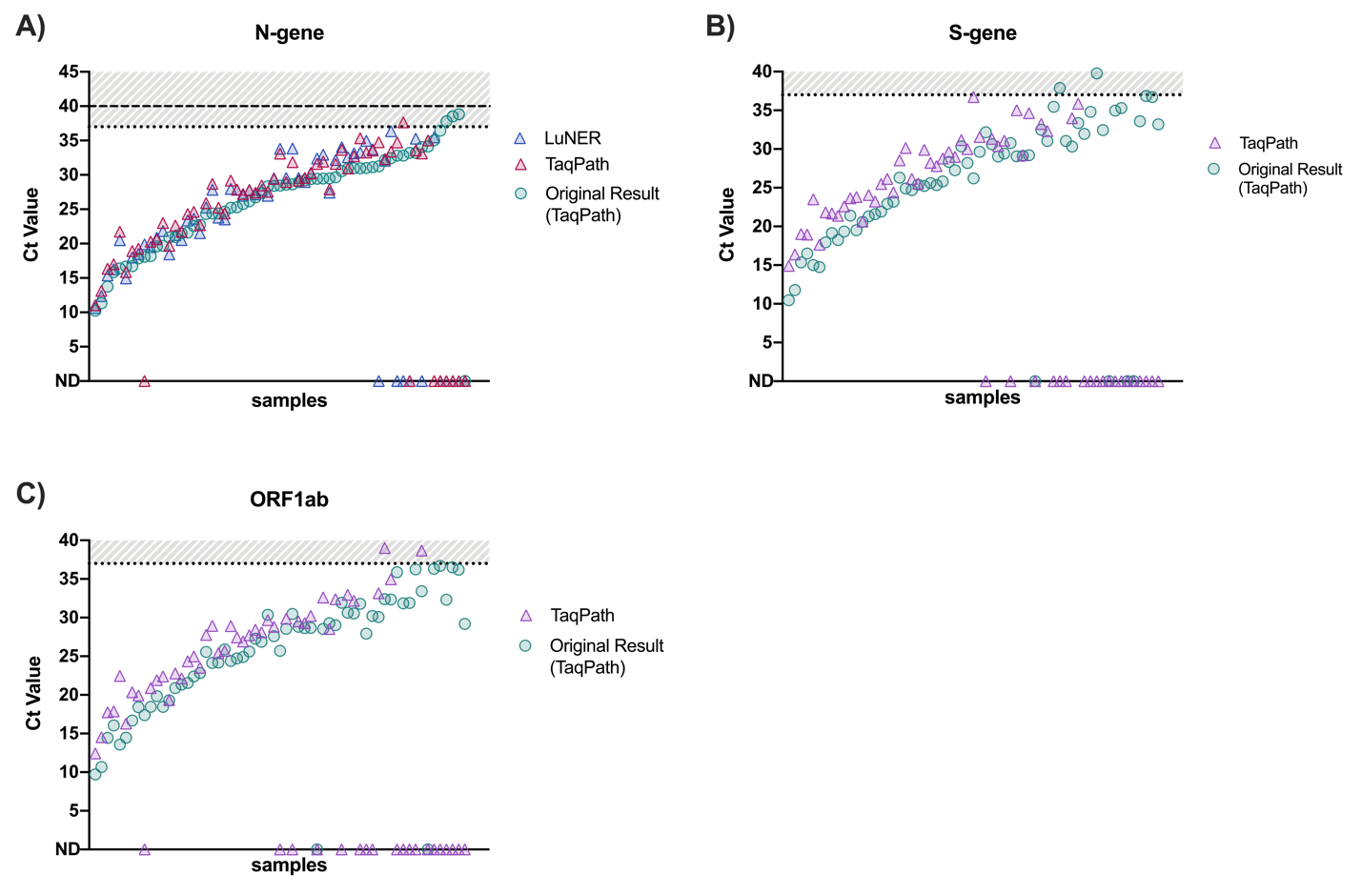

**Supplemental Figure 1.** Comparison of Ct values for 61 expected positive samples for A) N-gene, B) S-gene, and C) ORF1ab show that as the original Ct value increases, a greater number of undetected samples accumulate, suggesting loss of sensitivity upon re-testing.

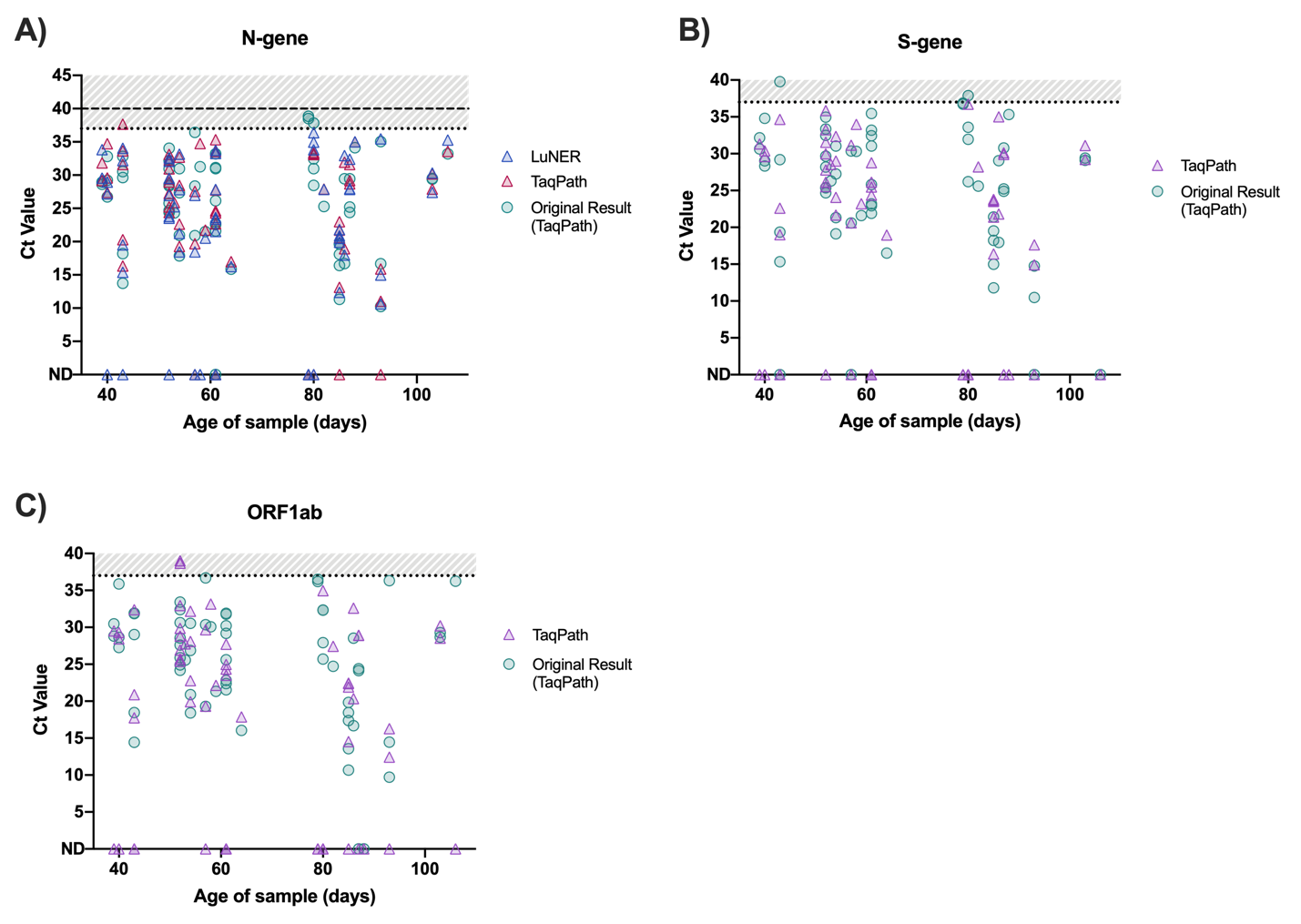

**Supplemental Figure 2.** Comparison of Ct values for 61 expected positive samples for A) N-gene, B) S-gene, and C) ORF1ab demonstrates that undetected samples were not correlated with receival date.
